# Investigating the Lived Experiences, Needs, and Stories (LENS) of Shwachman-Diamond Syndrome Patients and Caregivers

**DOI:** 10.64898/2026.11.14.26368095

**Authors:** Grace K. Lynch, Eszter S. Hars, Allison L. Cirino, Ashley S. Thompson, Vanessa L. Merker

## Abstract

**Background:** Shwachman-Diamond Syndrome is a rare genetic condition characterized by pancreatic insufficiency, skeletal differences, and hematologic manifestations. Previous research has focused on describing diagnosis and treatments of the disease. There is a gap in research highlighting SDS patient perspectives. This study aimed to elucidate the challenges of living with and seeking treatment for SDS based on the lived experiences of SDS patients and caregivers.

**Methods:** We conducted semi-structured interviews via Zoom or telephone calls with adults with SDS and caregivers of people with SDS. A semi-structured guide was developed by a multidisciplinary team and utilized for each interview. Interviews were transcribed verbatim and analyzed within the Dedoose platform. Three analysts performed inductive open coding of initial transcripts to collaboratively develop a codebook. The final codebook was applied to remaining transcripts by one analyst. Thematic analysis was performed to draw meaning from codes and identify common themes.

**Results:** 20 participants consented and were interviewed for this study, including 11 caregivers of loved ones with SDS and 9 adults with SDS. Two larger themes were identified: 1) SDS Impact on Daily Life and 2) Navigating Healthcare as an SDS Patient or Caregiver. Symptoms and management of SDS affected a wide range of areas, including careers, education, recreational activities, and socialization. When navigating the healthcare system, many participants shared that they felt misunderstood by providers and needed to frequently advocate for themselves or their loved ones. Numerous participants faced issues with accessing specialists who were knowledgeable or willing to learn about SDS and expressed the need for increased patient resources to help navigate a new diagnosis.

**Conclusions:** This study highlights the lived experiences of those impacted by SDS. Participants explained the impact of SDS symptoms and management on all aspects of daily life including career, socialization and extra-curricular activities. Additionally, people expressed that the development and distribution of SDS-specific resources are needed to improve both new patient and healthcare provider education. Increased awareness and education of the condition and the unique medical needs within clinical settings is necessary for expanding access to care.

## Background

Shwachman-Diamond Syndrome (SDS) is a rare disease that affects 1 in 75,000 live births in the United States (1). This condition is caused by biallelic pathogenic mutations in the Shwachman-Bodian-Diamond Syndrome (SBDS) gene in approximately 90% of cases (1,2). Less commonly, SDS or an SDS-like syndrome can also be caused by pathogenic biallelic mutations in the DNAJC21 and ELF1 genes, or heterozygous mutations in the SRP54 gene. SDS can have diverse clinical presentations and symptoms, including, but not limited to, exocrine pancreatic insufficiency, skeletal abnormalities, and hematologic manifestations, notably neutropenia and bone marrow failure (3). One of the more serious complications of SDS is the high risk for myelodysplastic syndrome (MDS) and acute myeloid leukemia (AML) with about 1 in 3 patients developing AML by the age of 30 years (1), and the risk continues to rise with age. Since its discovery in 1964, most research on SDS has focused on understanding the disease mechanisms and exploring treatment options rather than describing the lived patient experience.

Qualitative research can identify new concepts of interest missed in previous quantitative studies, contributing to a more holistic understanding of the patient experience (4–6). An understanding of patient needs is essential for optimizing future care, research, and advocacy initiatives to ultimately improve the quality of life for individuals affected by SDS. Research in other disease communities has allowed for communication of the complex experiences of patients and specific recommendations for change (5–7). For example, previous qualitative research on other inherited bone marrow failure syndromes (IBMFSs) has revealed insights into the patient experience beyond what is seen in clinic, such as increased anxiety about future risks, exhaustion with time-consuming treatment regimens, and changes made to patients’ future life plans (8–11). Additionally, many people with rare diseases express feeling pressured to be experts in their condition and emphasize the importance of connecting with the disease community for support (8).

One of the only studies exploring qualitative data from an SDS cohort utilized the Italian SDS registry. They distributed a questionnaire, consisting of open-ended, multiple choice and analogue scale questions, to analyze quality of life. They compared this to data from Cystic Fibrosis patients and found that SDS patients had increased rates of physical pain, cognitive impairment, and social struggles compared to Cystic Fibrosis patients (12). While this study provided insights into the unique physical and cognitive challenges SDS patients may face, the findings were limited due to the single country sample and scope of the questionnaire. A better understanding of holistic patient experiences, including the management of their care and psychological support needs, is necessary to improve care and research specific to Shwachman-Diamond Syndrome.

In recognition of the value of patient perspectives, the U.S. Food and Drug Administration (FDA) created patient-focused drug development meetings. These meetings were create with the goal of hearing directly from patients and caregivers, and to systematically gather patient experience data (13). Through these meetings, the FDA has gained access to a deeper understanding of specific disease burdens and has allowed patients to have a greater role in their own treatment development (13). SDS Alliance (SDSA) hosted an Externally-Led Patient-Focused Drug Development Meeting (EL-PFDD) in June 2025. The goal of this meeting was to provide patient perspectives to the FDA and those involved in SDS research and drug-development.

Understanding the SDS patient perspective through the SDS EL-PFDD meeting and associated research activities is critical to informing not only researchers’ and medical product developers’ insights into the unmet needs and risk tolerance of the SDS community, but also the FDA’s decisions during the drug development and approval processes.

This study was designed to expand on the information received at the EL-PFDD meeting. The meeting included specific stories shared in a short format from panels of patients and caregivers. The meeting and subsequent Voice of the Patient Report focused on understanding the specifics of people’s symptoms and experiences with current treatments (14). We aimed to capture the nuance of living with SDS through interviewing a wider range of participants and caregivers and giving them a platform to share stories in a less structured format. Through semi-structured qualitative interviews, we aimed to provide a space for patients and caregivers to share their experiences living with SDS. We focused on understanding the social and emotional impacts of SDS, including both the impact of the condition on day-to-day life and experiences navigating healthcare systems.

## Methods

This study was approved by the Mass General Brigham Institutional Review Board and the Northstar Review Board. Participants were recruited from the SDS Alliance Global Patient Survey and Collaboration Program (SDS-GPS), an established IRB-approved patient registry platform that collects patient-reported data through surveys and provides information about clinical trials and research to patients and families (15). Recruitment for SDS-GPS is conducted through emails, newsletters, social media posts, and presentations reaching approximately 200 families. Seventy participants are currently enrolled in this program and have consented under the Northstar IRB to be contacted about research opportunities. SDS-GPS program members were contacted by program staff via email and/or phone to invite them to participate in this qualitative study. Eligible participants were English-speaking individuals over 18 years of age who had either a personal diagnosis of SDS or who were caregivers for someone with SDS. A total of 29 eligible participants consented to the study; 9 were lost to follow-up during scheduling attempts, and 20 people completed the study. No compensation was provided for participation in this study.

Interviews followed a semi-structured interview guide developed by a multidisciplinary team with backgrounds in qualitative research, genetic counseling, and SDS. Questions covered a broad range of topics, including diagnostic journey, symptoms and treatments, experiences in healthcare, and hopes for future treatments. Participants were encouraged to share any notable challenges or experiences that they have had related to SDS. All interviews were conducted in English over Zoom between February 2025 and July 2025, each lasting between 30 to 70 minutes. All participants gave informed consent for study procedures, including recording of the interview.

Audio recordings of interviews were transcribed verbatim and analyzed using the Dedoose platform. Filler words, stutters, or other verbal markers that did not impact meaning have been removed from quotes for clarity. A team of 3 coders developed a codebook through an inductive, open-coding approach. The first third of transcripts were independently read by at least two coders, who did line-by-line coding to capture the meaning of each statement. The coding team then consolidated codes, collaboratively refined codebook definitions, and resolved any coding discrepancies between team members (16). Once the codebook was finalized through this process, a single coder applied codes to the remaining transcripts. The team met periodically to review coding throughout analysis in order to address any content with unclear meaning and ensure consistency in code application. Thematic analysis was then performed through grouping of similar codes and identification of underlying concepts and experiences captured within each code (17). Themes and subthemes were created to capture patterns in the data with exemplary quotes to represent each idea.

## Results

A total of 20 participants were interviewed (Table 1). Nine participants were adults with SDS, and 11 participants were caregivers of someone with SDS. Nine of the caregivers were parents of one child with SDS; one caregiver was a parent to two children with SDS, and one caregiver was a bereaved caregiver of a spouse with SDS. Twelve participants lived in the United States, and there was one participant each who lived in Australia, Canada, Ireland, Romania, Netherlands, Philippines, Poland and the United Kingdom.

**Table 1:** Participant Demographics.

|  | **Caregiver Participants n=11** | **Adults with SDS n=9** |
|---|---|---|
| **Location**<br>**(US;Other Countries)** | 5;6 | 7;2 |
| **Gender of affected individuals (Female; Male)** | 3;9 | 5;4 |
| **Median age of affected individual (Range)** | 3 years (1-44 years) | 43 years (22-49 years) |
| **Median age at diagnosis (Range)** | 5 months (5 days-43 years) | 23.5 years (1 month-40 years) |
| **Gender of caregiver (Female; Male)** | 5;6 | N/A |

Affected individuals had a wide range of symptoms. As outlined in Table 2, of the three key features of SDS, most participants reported pancreatic insufficiency (90%) and neutropenia (85%). Three of the adult participants had undergone bone marrow transplants.

**Table 2:**
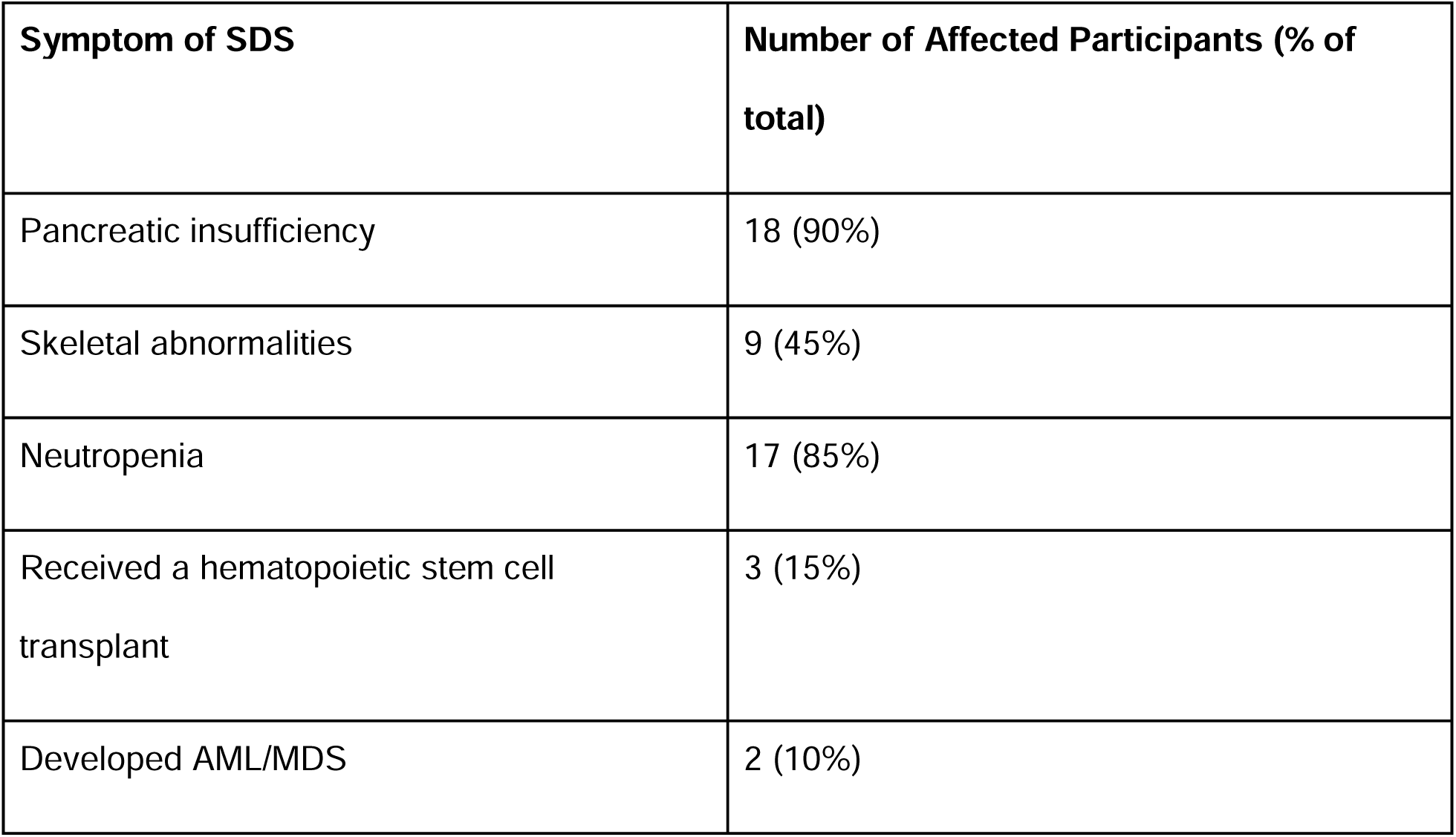
Symptoms of Affected Individuals (Adult Participants and Affected Loved Ones of Caregivers)

### Theme 1: SDS Impact on Daily Life

Participants described how numerous aspects of SDS, including management of the condition and treatment regimens, impacted their participation in various life activities, such as career development, extracurriculars, and socialization. This trend was seen in adult patients, pediatric patients, and caregivers of patients. As one adult patient explained:

> “… as I’ve learned, and as many other people with Shwachman-Diamond have learned, is that [SDS] doesn’t just affect your bone marrow. It affects all facets of your life.”

#### Inhibition of Career and Education Opportunities

Both caregivers and adults with SDS commonly described ways that SDS limited career development. Adults with SDS experienced challenges in holding the jobs that they want, frequently attributed to limited physical capacity to perform a job due to energy levels or physical pain. Others were limited by the need for decreased levels of social interactions in a work environment due to their compromised immune system. As a result, people were restricted in terms of the type of employment they could pursue. Many employed participants had to adjust the job type and/or amount of work they perform; as such, many sought positions that allowed for flexibility and accommodations.

> “Well, I have chronic pain and chronic fatigue. And it’s been difficult for me to have a normal job because I can’t really stand for long amounts of time. So luckily my job’s been really flexible. I do [take] a lot of breaks [with] a lot of sitting. I wouldn’t be able to do manual labor.”

Participants who were working often had to balance work with medical appointments. Many had to use their allotted paid time off for frequent medical appointments or hospitalizations. Even with a flexible position, one participant expressed struggles with taking so much time away from work:

> “I think my biggest challenge is my job. I have a lot of doctor appointments, and I go through my earned time really quickly. At the moment, I’m in negative earned time because I use all of it for appointments.”

Many caregivers of children living with SDS shared that they also had to adjust work schedules to accommodate the extensive care required for their children. One stay-at-home mom shared:

> “Our lifestyle is, schedule-wise, very demanding. [Child’s name] has a lot of appointments, and if I had a regular [full-time] job, it would be really hard for me to be able to take him to all of his appointments.”

Other parents expressed how, even with accommodating employers, they were still limited in their opportunities for career growth. One participant shared the largest impact on her personal life as:

> “I would say a delay of career advancement. I have a fantastic boss. He gives me all this time off I could possibly need for my son, and flexible schedule and all that stuff. But I’ve been passed over for several promotions for people who definitely have less knowledge than I do, [and/or] been with the company less [time] than I have.”

Other participants expressed a similar sentiment about absences from educational opportunities. Caregivers of children with SDS discussed needing to take their child out of school often. They felt this impacted their child both academically and socially. Despite these challenges in maintaining consistent attendance in primary and high school, all adult patients in our sample pursued higher education. They often had to consider many factors when deciding where to go, such as staying near home or near adequate medical facilities to accommodate consistent appointments and potential hospitalization.

#### Ability to Participate in Recreational Activities

Many affected adults highlighted their limited ability to participate in sports or physical activities. People were impacted by fatigue, physical pain, and/or exacerbation of symptoms during exercise. Those who underwent hematopoietic stem cell transplantation reported interrupted activities for an extended period of time such as the inability or limited ability to work, attend school, and/or simply leave their homes or the hospital. Participants expressed frustration at “missing out” on social or personal activities that they wanted to participate in but were prevented from doing so due to their physical limitations.

“I think just in general, I’m not normal anymore, I mean, I don’t have the energy. Before [my SDS diagnosis], I was very active in sports and outdoor activities. I can’t do any of that at the moment. I don’t have the energy for it. Simple stuff like mowing my lawn just drains me.”

People often made changes to their daily routines and social schedules in order to accommodate decreased energy levels. As one bereaved caregiver of an affected adult shared:

> “It’s frustrating, you know, when you can see other people are going out for work five days a week and still socialize, and she just almost had to kind of learn and force herself to just balance things really carefully.”

Caregivers outlined the ways that their child’s play was also impacted by their physical and social capacities. Many caregivers took precautions to minimize exposure to pathogens due to their child’s compromised immune system, such as avoiding high-touch, indoor environments (e.g., playgrounds and daycares). As a result, caregivers expressed concern that their children missed out on opportunities for play and socialization.

> “Right now, I have to—he probably can’t—I don’t take him to any indoor play zones or play lands. And he would love it. He’s super social…some of his social opportunities and opportunities for other children are limited and hindered [because of SDS].”

The symptoms and treatments of SDS also impacted the ability of both individuals and families to travel. Patients and caregivers had to consider daily treatment needs, health risks, and access to care before planning any trips. In many cases, participants were cautious about leaving their trusted clinicians for an extended period of time. Some participants reported planning vacations around locations with nearby hospitals that could manage any potential emergencies. This is one example of a change to lifestyle that many families had to make following a diagnosis of SDS.

“I messaged [the doctor] about it, and I said—because you have to have a yellow fever jab—inoculation or immunization—on your passport, otherwise you’re not allowed in. And I asked him about it. I said, ‘is it safe for me to have this?’ And he said no.”

#### Feelings of Loneliness

Participants described how barriers to socializing contributed to feelings of loneliness. Adult participants reflected on past feelings of isolation from classmates in primary school due to their condition. Caregivers of young patients similarly expressed concerns about their child feeling different or left out. Affected individuals described feeling “othered” by classmates who didn’t understand why they were sick or missing class for appointments. As one adult participant with SDS described:

“I do remember spending a lot of time in the hospital and then in the [healthcare facility] and then at home recovering. I am very glad that the school that I went to was very supportive and very understanding. Even though, you know, I was a kid, I felt isolated from my friends, I did feel a tad lonely at times.”

Having a condition that the general population did not know about also made many adult patients feel more emotionally isolated. In many cases, people were hesitant to open up to friends about what they were going through because they did not have the energy to explain SDS or thought that people would not understand. Having experiences that greatly contrasted with their peers’ made people feel less known. As one participant shared:

“Having that constant feeling of being different, it can definitely take a toll on your mental health and make you feel either emotionally strained or lonely at times.”

### Theme 2: Navigating Healthcare as an SDS Patient or Caregiver

People also expressed a wide range of challenges navigating the healthcare system as they sought to diagnose and treat their/their loved one’s SDS symptoms. These experiences often took an emotional toll on participants. Most patients and caregivers were heavily involved in aspects of the medical management of SDS, including advocating for necessary procedures, seeking insurance coverage, and educating providers about SDS. Many participants expressed that they felt they would not have received the same level of care if they had not been active advocates for themselves or their loved ones.

#### Understanding an SDS Diagnosis

Upon receiving a diagnosis, many families reported being given a brief description of SDS, but most did not have extensive conversations with providers about what the future could look like. Because of this, many participants reported conducting independent research on SDS. Most individuals turned to the internet as their primary source of information. This led to some helpful outcomes, such as finding simplified information on SDS or connecting with SDS organizations. However, participants also felt responsible for sorting through the extensive information they found on the internet to learn about SDS. In numerous cases, participants faced barriers to accessing information due to paywalls on peer-reviewed research, with some paying out of pocket to access articles. Participants described increased anxiety with the amount of information they found when trying to research and understand SDS.

> “When you go on the Google search, and there’s a bunch of stuff you see that does cause—create a lot of fear. So, it was a lot of fear in the beginning.”

Many people turned to online SDS communities through Facebook or other various organizations for information about SDS and others’ direct experiences with it. Since the symptoms and severity of those symptoms present on such a large spectrum in patients with SDS, many participants reported feeling overwhelmed with the possible outcomes for themselves or their child. They often reported reading stories of other SDS-impacted families and described not knowing how to make sense of the large range of cases described online. One participant explained her experience reading social media posts about other children with SDS:

> “ I think it just panicked me even more or kind of, you know, drilled into my brain that all these things that happened to her were going to happen to my son.”

At the same time, affected individuals and their families frequently found community and reassurance through online groups of people and families affected by SDS. Often, there was a sense of validation as participants commonly reported never meeting someone else with SDS prior to finding an online group. This fostered a strong community of people looking to provide hope and support to each other.

> “It’s actually really cool because there are a few really close friends of mine who have [Shwachman-Diamond syndrome] who I’ve been able to reach out to and talk about my syndrome with. So, they’ve been really helpful in either comforting me or just staying in communication just so I know that, hey, I don’t have to go through this alone.”

Since SDS is commonly diagnosed in childhood, caregivers also have to navigate establishing an understanding of SDS for both their affected and unaffected children. Caregivers expressed struggles in determining how much information to tell their children at different ages. Adult patients expressed that many of them did not understand the diagnosis fully until their adult years.

> “It’s not really until…after I went to university that I truly understood what [SDS] was because there was a consultant hematologist that sat me down and went exactly through it. They didn’t skirt around the issues of it. They laid it all on the line for me, and I was like, actually, that’s quite helpful, really.”

#### Impact of Providers’ Knowledge of SDS

Participants described feeling that many healthcare professionals were not knowledgeable about SDS. This impacted the patient experience in all stages of care beginning with the process of receiving a diagnosis. Many participants underwent a diagnostic odyssey in which multiple providers went through a lengthy process of ruling out other diagnoses before SDS. In some cases, parents were the ones to mention SDS as a possible diagnosis after conducting their own research on the symptoms they were seeing in their child. Caregivers described the need to be proactive in requesting specific tests for a potential SDS diagnosis and exploring treatment options even without a genetic diagnosis to improve quality of life.

“We started immediately looking, learning about Shwachman-Diamond [syndrome]. As a matter of fact, the gastroenterologist wasn’t going to put him on [pancreatic enzyme replacement therapy] until they got the genetic confirmation. We convinced [his team] to go ahead and put him on the enzymes even without that, while we were waiting for that one month.”

Many patients and caregivers recounted experiences with doctors who had never heard of SDS. Participants commonly felt like the “expert in the room” and faced the burden of educating healthcare professionals on the condition, which, in some cases, led to mistrust when receiving care from doctors who admitted they only recently learned what SDS was.

> “It’s been really difficult because I just got a new [primary care physician (PCP)], and my new PCP doesn’t know anything about [Shwachman-Diamond syndrome], so I’m trying to educate her. Like every doctor I go to, I have to educate them because they don’t know anything. Like they’ve never heard of [SDS].”

As a result, many individuals described becoming strong advocates for themselves or their children. They learned how to quickly educate medical providers on the most serious aspects of SDS and their care. Participants expressed the importance of being trusted and believed by providers, as they sometimes felt like they were questioned when expressing their experiences and needs. People often had to strongly push for their specific needs in emergency situations.

> “Having to explain [SDS] and having to advocate to say, ‘I need to get a room. I am not able to sit in the waiting room for too long. I cannot risk being exposed to this and that’ I think that was definitely an obstacle, a challenge.”

In some cases, the lack of knowledge about SDS from medical providers led to denial of care. Participants shared situations where a medical center did not want to admit or treat them due to providers’ lack of familiarity with their condition. Often, people relied on a doctor who was knowledgeable about SDS to help advocate for them in these situations.

> “Being turned away from cancer treatment because [the providers] were scared…that was kind of alarming. I was like, ‘What do you mean? I have to go all the way to [a distant hospital] and leave my family and leave my kids?’ It was really hard. When rather, they should just say ‘Oh, they’re doctors, they should research about my syndrome instead of just turning me away.’”

In contrast, when providers dedicated time to researching and understanding SDS, participants had more positive experiences. Many people who were receiving care at larger medical centers interacted with doctors who had more experience with SDS. In some cases, doctors were not familiar with SDS but expressed a commitment to learning about the condition alongside the patient and serving as a resource for them. Participants often reported feeling more supported by their care team when they could rely on their providers to answer questions or provide additional information.

> “Once the doctors figure out what [SDS] is, a lot of the doctors that I’ve worked with…were willing to do the research, willing to take the time to learn, and willing to explore more into this area.”

Adults with SDS expressed additional struggles navigating care in their transition from pediatric to adult care, particularly in finding and educating doctors who would treat them. This barrier to care was exacerbated by the lack of research and knowledge on adults with SDS.

> “It seems like when you do become an adult with [Shwachman-Diamond syndrome], it’s very easy for you to almost fall off the map, like fall through the hole.”

#### Barriers to Care

In the United States (US), insurance was highlighted as one of the main barriers to accessing SDS-related care and medications. Participants described that one of the driving influences on this issue was the lack of recognition of SDS as a specific disease entity by insurance companies. With SDS only receiving an official ICD-10-CM code in the US in 2023, many people reported having coverage denied because insurance companies were unfamiliar with SDS and did not consider treatments medically necessary.

In countries outside of the US, participants described facing similar barriers to care, as SDS was sometimes not nationally recognized as a disease. In one case, a family was required to internationally ship their child’s pancreatic enzyme replacement therapy since this medication was not available in their home country, requiring additional effort and money. Some international participants also highlighted that lack of coverage seemed to result from insurance companies’ unfamiliarity with the condition.

> “[SDS] is not recognized because say if you have cystic fibrosis, which is the nearest comparable thing, people who have that diagnosis get their [pancreatic enzyme replacement therapy medication]. They’re exempt from paying.”

In some cases, the treatment itself presented barriers. Many caregivers of children with SDS faced difficulties in ensuring their child took the proper medications. Whether it was timing the administration of pancreatic enzyme replacement therapy capsules with their child’s mealtimes or trying to get their child to swallow pills, parents reported having to find creative solutions, often leading to an increased physical and emotional toll on the caregivers.

> “Every doctor we talked to wanted to give us another suggestion of how to get [his medication] down his throat. We tried everything, and it felt like we were tormenting our child all the time, and nobody could understand how difficult that was on a daily basis.”

#### Navigating Multiple Providers and Specialties

People had differing experiences navigating care across multiple specialties. When providers were flexible in scheduling appointments and communicated promptly with other specialists, participants expressed an increased sense of support. People also felt more reassured when their providers were aware of and agreed with their care plan.

In other cases, participants described being responsible for communication between providers to schedule and coordinate care. People reported feeling obligated to confirm with multiple providers that the treatments they, or their child, were receiving were the safest and best option, placing additional burden on participants to communicate their/their child’s symptoms and care plan to all providers. As one parent shared:

> “A lot of advocating and fighting for him, and pushing to get him the best therapies and services, and making sure I’m happy with all the doctors in his care. It’s tough. It’s like a full-time secretary job”

In some cases, patient intervention was necessary for coordinating appropriate care, particularly if patients felt like their specialists weren’t considering the different body systems impacted by SDS. One mother explained their experience coordinating a procedure in one department while ensuring they follow guidelines set by another doctor. She explained how her intervention in coordinating care impacted management :

> “I’m like, ‘Wait a minute, hold on. We couldn’t use a rectal thermometer on our son because of the potential for an infection. And you want to do what now?’ So luckily, we had a hematology oncology appointment the next week… she said, ‘Oh, we can do this [procedure], but he needs to go on the granulocyte colony-stimulating factor (GCSF)] to boost his bone marrow before he has [a procedure]’…the GI doctor would never have done that. He didn’t think of that.”

## Discussion

This qualitative study explored experiences of people in the Shwachman-Diamond syndrome community, specifically outlining the impact of an SDS diagnosis on patients’ and caregivers’ daily lives and the difficulty of navigating the healthcare system as an SDS patient or caregiver. Participants highlighted how SDS symptoms and the extensive effort required to manage the condition influenced many aspects of their lives, including work/school, socialization, and leisure activities.

Participants reported facing multiple forms of social isolation through both limitations in their ability to participate in social activities and feelings of alienation from people who did not have shared experiences or understanding. In many cases, people connected with SDS community groups through social media or advocacy organizations to address this social isolation and gain knowledge about the condition. These groups gave participants a forum to ask questions and learn about people’s direct experiences, yet the wide range of clinical presentations of SDS often could cause increased anxiety and uncertainty in participants. Previous research has shown that these feelings are echoed by people in other rare disease communities. Caregivers and families of those with a rare disease have expressed feeling misunderstood by their friends and families who did not share similar experiences (18–20). Rare disease groups, often found through online settings, can provide a source of comfort and support from people who have had similar experiences (21,22). At the same time, research has shown how online groups can require people to spend time sorting through what information is relevant to them and ensuring what they are accessing is reliable (22). Along with this, our research shows how online communities may also increase anxiety when people are seeing a large range of the possible impacts and outcomes of a disease.

Overall, our participants found self-advocacy necessary to find providers who are familiar with SDS or educate those who were unfamiliar about the condition. Qualitative research on other rare diseases has similarly reported that many aspects of patients’ and caregivers’ lives, including career, education and socialization, are impacted by the symptoms and management of their condition (8,10,20). In a systematic literature review comparing experiences of adults with various rare diseases, similar themes were found with difficulties navigating the healthcare system (23). Not unique to the SDS community, many rare disease patients have also reported feeling their care is impacted by the lack of knowledge about their condition in the medical field (23,24). The SDS participants reported how this also impacted insurance coverage and medication availability, as SDS only recently received an ICD-10 code in the US in 2023 (25). Additionally, research within the rare disease community has highlighted experiences of “expert patients” who are tasked with researching and understanding their condition and subsequently explaining it to providers (23,26,27). These common experiences highlight the need for broader changes in how care is provided to those affected by rare disease (28,29).

### Clinical Implications

As many of the challenges faced by SDS patients and caregivers were related to the lack of providers knowledgeable about SDS, increased awareness of SDS in the medical field and more resources for providers to quickly and easily understand the condition could greatly benefit SDS families. One method that has been implemented in the United Kingdom to aid in this process is a patient passport (26,30). This document, developed through collaboration between providers and patients, is given to new providers as a quick outline of the most important information about a patient and their rare condition. This resource may incorporate information from the most up-to-date consensus guidelines for SDS while focusing on the specific patient’s symptoms and treatments (31). A similar resource has been developed for SDS to help kids in the school setting better explain their diagnosis to teachers and classmates (32). Applying this resource in the medical setting may be especially beneficial for people with SDS, as the condition has a wide range of symptoms and severity that impact each patient differently. Research shows that implementing this type of resource can benefit both providers and patients in communicating care history and needs (26).

Often, participants were put in the position of ensuring that the care they were receiving across different hospitals and/or specialties was aligned. Regular meetings between the healthcare team and collaboration with experts could enhance care coordination and provide space to discuss ways to improve various treatment plans. Patient care navigators and social workers also serve as a resource to aid in the delivery of care. Research has shown that the involvement of primary care providers (PCPs) in coordination of care has been beneficial to those with complex care needs (33). In other areas of disease, such as oncology, multidisciplinary frameworks have been used to improve care such as through multidisciplinary programs or conferences (34–36). Additionally, the integration of holistic care pathways has been used to facilitate rare disease care in many healthcare systems, primarily in Europe (37–39). Care pathways develop evidence-based care recommendations through consensus guidelines and national expertise, and translate them into the context of local healthcare systems and patient-specific needs (35). While care pathways require integration at multiple levels, they have been shown to be beneficial to patients with conditions that require high levels of care coordination (38,39). Implementation of innovative approaches to care coordination for SDS patients could release the burdens of navigating care from patients and caregivers, ultimately reducing their stress and the level of time required to keep up with care management.

### Study Limitations and Future Directions

As recruitment occurred solely through SDS Alliance’s SDS-GPS study platform, all participants were actively involved in this community and seeking opportunities to participate in future research. The experiences of individuals who haven’t had the capacity to access care or seek out patient communities might differ from the reports of our participants. Additionally, as we only conducted interviews in English, there was limited capacity to understand the full range of experiences for international and non-English-speaking participants. However, through the inclusion of participants from nine different countries, we were able to identify and describe shared experiences of SDS across various healthcare systems. Future research with international participants could identify healthcare-system-specific vulnerabilities and strengths to support optimal implementation of clinical programs to improve SDS care. Additionally, our inclusion criteria for participants was limited to caregivers and adults with SDS. As many patients with SDS are children, assessing the perspectives of pediatric patients could provide more insight into the understanding of SDS and its impact on lives in childhood. Previous research focused on the views of children with rare diseases has been able to provide insights into their feelings and day-to-day experiences that may not be captured as fully through interviews with caregivers (40).

Finally, our analysis focused specifically on the healthcare journeys of people with SDS and the impact that managing SDS symptoms and healthcare has on patients’ and caregivers’ daily lives rather than exploring all aspects of SDS in depth. However, additional details on the range of symptoms experienced by SDS patients and patient/caregivers’ perspectives on current and future SDS treatments are covered extensively in the Voice of the Patient report summarizing a recent Externally Led Patient Focused Drug Development on SDS (14). Areas that were briefly discussed which may be explored further in future research are specific experiences within SDS management, such as those who underwent stem-cell transplant or those who have transitioned from pediatric to adult care.

## Conclusions

In recent years, there has been an increased emphasis on learning directly from patients about their experiences with rare disease and hopes for future change (13,41,42). This study provides a foundation for understanding the lived experiences of people with SDS and highlights the various ways that care can be improved to better support this community. Future research can focus on gaining a deeper understanding of specific experiences within the SDS community, such as experiences with a hematopoietic stem cell transplant, the transition from pediatric to adult care, and the cancer journey of those who developed MDS, AML, and/or another malignancy. SDS families face numerous challenges living with SDS and navigating care, but collaborative efforts from providers, healthcare systems, and patient advocacy groups to improve awareness, coordinate care, and innovate treatments hold the promise of beginning to address their needs.

## Data Availability

The dataset generated during the current study are not publicly available due to privacy/ethical restrictions.

## List of Abbreviations

AML: Acute myeloid leukemia
FDA: U.S. Food and Drug Administration
GCSF: granulocyte colony-stimulating factor
IBMFS: Inherited bone marrow failure syndrome
MDS: Myelodysplastic syndrome
PCP: Primary care provider
SBDS: Shwachman-Bodian Diamond Syndrome
SDS: Shwachman-Diamond Syndrome
SDSA: Shwachman-Diamond Syndrome Alliance
SDS-GPS: SDS Alliance Global Patient Survey and Collaboration
Program US: United States

## Declarations

### Ethics approval and consent to participate

Approval was granted by the Mass General Brigham Institutional Review Board (Protocol #2024P003441) and the Northstar Review Board. All participants provided verbal consent to participate in this study. Study procedures were performed in accordance with the Belmont Report and U.S. Common Rule 45 CFR 46.

### Clinical trial number

Not applicable.

### Consent for publication

Not Applicable

### Authors’ contributions

AST, EH, and VLM conceived of the work, and all authors help design the study. GL, AST, EH, and VLM developed the semi-structured interview guide. EH recruited participants. GL and AST conducted the qualitative interviews. GL, AST, and VLM coded the data and analyzed transcripts. GL drafted the manuscript, and EH, AC, AST, and VLM revised the manuscript for important intellectual content. All authors reviewed and approved the final version to be published.

## References

1. Kawashima N, Oyarbide U, Cipolli M, Bezzerri V, Corey SJ. Shwachman-Diamond syndromes: clinical, genetic, and biochemical insights from the rare variants. Haematologica. 2023 May 25;108(10):10. doi:10.3324/haematol.2023.282949

2. Nelson A, Myers K. Shwachman-Diamond Syndrome. In: Adam MP, Feldman J, Mirzaa GM, Pagon RA, Wallace SE, Amemiya A, editors. GeneReviews® [Internet]. Seattle (WA): University of Washington, Seattle; 1993 [cited 2024 Nov 24]. Available from: http://www.ncbi.nlm.nih.gov/books/NBK1756/ PubMed PMID: 20301722.

3. Farooqui SM, Ward R, Aziz M. Shwachman-Diamond Syndrome. In: StatPearls [Internet]. Treasure Island (FL): StatPearls Publishing; 2025 [cited 2025 Jan 16]. Available from: http://www.ncbi.nlm.nih.gov/books/NBK507866/ PubMed PMID: 29939643.

4. Deal LS, Goldsmith JC, Martin S, Barbier AJ, Roberds SL, Schubert DH. Patient Voice in Rare Disease Drug Development and Endpoints. Ther Innov Regul Sci. 2017 Mar 1;51(2):257–63. doi:10.1177/2168479016671559

5. Lyons AC. Advancing and extending qualitative research in health psychology. Health Psychol Rev. 2011 Mar 1;5(1):1–8. doi:10.1080/17437199.2010.544638

6. Marzban S, Najafi M, Agolli A, Ashrafi E. Impact of Patient Engagement on Healthcare Quality: A Scoping Review. J Patient Exp. 2022 Sep 16;9:23743735221125439. doi:10.1177/23743735221125439 PubMed PMID: 36134145; PubMed Central PMCID: PMC9483965.

7. Smits RM, Vissers E, Te Pas R, Roebbers N, Feitz WFJ, van Rooij IALM, et al. Common needs in uncommon conditions: a qualitative study to explore the need for care in pediatric patients with rare diseases. Orphanet J Rare Dis. 2022 Apr 4;17(1):153. doi:10.1186/s13023-022-02305-w PubMed PMID: 35379257; PubMed Central PMCID: PMC8981675.

8. Bogart KR, Limon M, Fischer S, Voss ME. “I would love to talk to someone that actually understands”: Psychosocial experiences of adults with Fanconi anemia. J Health Psychol. 2025 Aug 3;13591053251353200. doi:10.1177/13591053251353200

9. Knudsen KB, Boisen KA, Katzenstein TL, Mortensen LH, Pressler T, Skov M, et al. Living with cystic fibrosis – a qualitative study of a life coaching intervention. Patient Prefer Adherence. 2018 Apr 19;12:585–94. doi:10.2147/PPA.S159306

10. Liu T, Pan Y, Ye M, Sun Q, Ding X, Xu M. Experience of life quality from patients with aplastic anemia: a descriptive qualitative study. Orphanet J Rare Dis. 2023 Dec 21;18(1):393. doi:10.1186/s13023-023-02993-y

11. Zierhut HA, Bartels DM. Waiting for the next Shoe to Drop: The Experience of Parents of Children with Fanconi Anemia. J Genet Couns. 2012;21(1):45–58. doi:10.1007/s10897-011-9394-5

12. Perobelli S, Nicolis E, Assael BM, Cipolli M. Further characterization of Shwachman-Diamond syndrome: psychological functioning and quality of life in adult and young patients. Am J Med Genet A. 2012 Mar;158A(3):567–73. doi:10.1002/ajmg.a.35211 PubMed PMID: 22315206.

13. Chalasani M, Vaidya P, Mullin T. Enhancing the incorporation of the patient’s voice in drug development and evaluation. Res Involv Engagem. 2018 Apr 2;4(1):10. doi:10.1186/s40900-018-0093-3

14. Hars ES, Merker VL. Voice of the Patient Report for Shwachman-Diamond Syndrome [Internet]. 2026 May 11. doi:10.5281/zenodo.20126868

15. SDS Alliance [Internet]. [cited 2025 Dec 9]. SDS-GPS: Make your voice count | Shwachman-Diamond Syndrome Alliance. Available from: https://www.sdsalliance.org/sds-gps

16. Hemmler VL, Kenney AW, Langley SD, Callahan CM, Gubbins EJ, Holder S. Beyond a coefficient: an interactive process for achieving inter-rater consistency in qualitative coding. Qual Res. 2022 Apr 1;22(2):194–219. doi:10.1177/1468794120976072

17. Guest G, MacQueen K, Namey EE. Applied Thematic Analysis [Internet]. SAGE Publications; 2012 [cited 2026 Oct 2]. 320 p. Available from: https://www.sagepub.com/shop/buy-a-book/applied-thematic-analysis-1-233379

18. Currie G, Szabo J. Social isolation and exclusion: the parents’ experience of caring for children with rare neurodevelopmental disorders. Int J Qual Stud Health Well-Being. 2020 Jan 1;15(1):1725362. doi:10.1080/17482631.2020.1725362 PubMed PMID: 32048917.

19. Belzer LT, Wright SM, Goodwin EJ, Singh A, Carter BS. Psychosocial Considerations for the Child with Rare Disease: A Review with Recommendations and Calls to Action. Children. 2022 Jun 20;9(7). doi:10.3390/children9070933

20. Wilsnack C, Rising CJ, Pearce EE, Forbes Shepherd R, Thompson AS, Majid A, et al. Defining the complex needs of families with rare diseases—the example of telomere biology disorders. Eur J Hum Genet. 2024 Dec;32(12):1615–23. doi:10.1038/s41431-024-01697-6

21. Pearce E, Raj H, Emezienna N, Gilkey MB, Lazard AJ, Ribisl KM, et al. The Use of Social Media to Express and Manage Medical Uncertainty in Dyskeratosis Congenita: Content Analysis. JMIR Infodemiology. 2024 Jan 15;4:e46693. doi:10.2196/46693 PubMed PMID: 38224480; PubMed Central PMCID: PMC10825764.

22. Ashtari S, Taylor A. Patients With Rare Diseases and the Power of Online Support Groups: Implications for the Medical Community. JMIR Form Res. 2023 Sep 14;7:e41610. doi:10.2196/41610 PubMed PMID: 37707878; PubMed Central PMCID: PMC10540027.

23. von der Lippe C, Diesen PS, Feragen KB. Living with a rare disorder: a systematic review of the qualitative literature. Mol Genet Genomic Med. 2017;5(6):758–73. doi:10.1002/mgg3.315

24. Mease C, Fermaglich LJ, Jackler K, Shermer S, Miller KL. Determining Commonalities in the Experiences of Patients with Rare Diseases: A Qualitative Analysis of US Food and Drug Administration Patient Engagement Sessions. The patient. 2024 Jan;17(1):25–37. doi:10.1007/s40271-023-00648-5 PubMed PMID: 37833521; PubMed Central PMCID: PMC11841686.

25. Hars ES, McReynolds LJ. From Challenge to Opportunity: How Shwachman-Diamond Syndrome Became a Promising Target for Therapy Development. Clin Pharmacol Ther. 2024 Dec;116(6):1377–80. doi:10.1002/cpt.3393 PubMed PMID: 39039619; PubMed Central PMCID: PMC11567794.

26. Balfour J, Morrison V, Seed L, Clymer J, Warnants E, Lampkin A, et al. Patient passports for rare diseases: results of a pilot study. Eur J Hum Genet. 2026 Jan;34(1):99–107. doi:10.1038/s41431-025-01930-w

27. Budych K, Helms TM, Schultz C. How do patients with rare diseases experience the medical encounter? Exploring role behavior and its impact on patient-physician interaction. Health Policy. 2012 May;105(2–3):154–64. doi:10.1016/j.healthpol.2012.02.018 PubMed PMID: 22464590.

28. Kole A, Hedley V. Recommendations from the Rare 2030 Foresight Study: The future of rare diseases starts today. https://eprints.ncl.ac.uk [Internet]. 2021 [cited 2026 Mar 9]. Available from: https://eprints.ncl.ac.uk

29. Stoller JK. The Challenge of Rare Diseases. CHEST. 2018 Jun 1;153(6):1309–14. doi:10.1016/j.chest.2017.12.018 PubMed PMID: 29325986.

30. Lee LK, Mulvaney-Day N, Berger AM, Bhaumik U, Nguyen HT, Ward VL. The Patient Passport Program: An Intervention to Improve Patient–Provider Communication for Hospitalized Minority Children and Their Families. Acad Pediatr. 2016 Jul 1;16(5):460–7. doi:10.1016/j.acap.2015.12.008 PubMed PMID: 26724179.

31. Dror Y, Donadieu J, Koglmeier J, Dodge J, Toiviainen-Salo S, Makitie O, et al. Draft consensus guidelines for diagnosis and treatment of Shwachman-Diamond syndrome. Ann N Y Acad Sci. 2011;1242(1):40–55. doi:10.1111/j.1749-6632.2011.06349.x

32. Hars ES, Shwachman-Diamond Syndrome Alliance. Personalized School Letter Tool for Shwachman-Diamond Syndrome (SDS) [Internet]. Zenodo; 2026 [cited 2026 Sep 15]. Available from: https://zenodo.org/doi/10.5281/zenodo.21324915 doi:10.5281/ZENODO.21324915

33. Vidic N, McGlynn A, Abdi F, Tam CWM, Crampton RM, Lim KS, et al. Integrated Care for People Living With Rare Disease: A Scoping Review on Primary Care Models in Organization for Economic Cooperation and Development Countries. J Prim Care Community Health. 2025 Oct 1;16:21501319241311567. doi:10.1177/21501319241311567

34. Meguid C, Ryan CE, Edil BH, Schulick RD, Gajdos C, Boniface M, et al. Establishing a framework for building multidisciplinary programs. J Multidiscip Healthc. 2015 Dec 2;8:519–26. doi:10.2147/JMDH.S96415 PubMed PMID: 26664132; PubMed Central PMCID: PMC4671763.

35. Pawlik TM, Laheru D, Hruban RH, Coleman J, Wolfgang CL, Campbell K, et al. Evaluating the Impact of a Single-Day Multidisciplinary Clinic on the Management of Pancreatic Cancer. Ann Surg Oncol. 2008 Aug;15(8):2081–8. doi:10.1245/s10434-008-9929-7 PubMed PMID: 18461404; PubMed Central PMCID: PMC2907242.

36. Wright FC, Vito CD, Langer B, Hunter A. Multidisciplinary cancer conferences: A systematic review and development of practice standards. Eur J Cancer. 2007 Apr 1;43(6):1002–10. doi:10.1016/j.ejca.2007.01.025

37. Castro R, Senecat J, de Chalendar M, Vajda I, Dan D, Boncz B. Bridging the Gap between Health and Social Care for Rare Diseases: Key Issues and Innovative Solutions. In: Posada de la Paz M, Taruscio D, Groft SC, editors. Rare Diseases Epidemiology: Update and Overview [Internet]. Cham: Springer International Publishing; 2017 [cited 2026 Apr 6]. p. 605–27. Available from: https://doi.org/10.1007/978-3-319-67144-4_32 doi:10.1007/978-3-319-67144-4_32

38. Chou C, Wiredu SO, Von Imhof L, Tan A, Agarwal S, Lydston M, et al. Health services interventions to improve the quality of care in rare disease: a scoping review. Ther Adv Rare Dis. 2024 May 1;5:26330040241262810. doi:10.1177/26330040241262810

39. Tumiene B, Graessner H. Rare disease care pathways in the EU: from odysseys and labyrinths towards highways. J Community Genet. 2021 Apr 1;12(2):231–9. doi:10.1007/s12687-021-00520-9

40. Buckle N, Rogers Y, O’Toole D, McNulty S, Kroll T, Gibbs L, et al. A qualitative exploration of children’s lives with rare diseases. Child Care Health Dev. 2024;50(4):e13294. doi:10.1111/cch.13294

41. Quintal A, Carreau I, Grenier AD, Hébert C, Yergeau C, Berthiaume Y, et al. An Ethics Action Plan for Rare Disease Care: Participatory Action Research Approach. J Particip Med. 2023 Nov 23;15(1):e46607. doi:10.2196/46607

42. Velvin G, Hartman T, Bathen T. Patient involvement in rare diseases research: a scoping review of the literature and mixed method evaluation of Norwegian researchers’ experiences and perceptions. Orphanet J Rare Dis. 2022 May 31;17(1):212. doi:10.1186/s13023-022-02357-y

